# Overground suspension walkers elicit more and higher quality stepping than treadmills: A longitudinal study of pre-walking infants with Down syndrome

**DOI:** 10.64898/2026.05.05.26352150

**Authors:** Christina M. Hospodar, Florencia A. Enriques, Aylin I. Paez, Heather A. Feldner, Julia E. Looper, Kari S. Kretch

**Affiliations:** University of Southern California; University of Washington; University of Puget Sound

## Abstract

**Purpose:** Children with Down syndrome (DS), a genetic condition associated with neuromotor impairments, walk ∼1 year later than typically developing peers. To inform future interventions, we measured infants’ stepping behaviors in a novel task—overground stepping in a suspension walker—compared to supported treadmill stepping.

**Materials and Methods:** We tested 31 pre-walking infants with Down syndrome across 71 sessions at 10, 13, 16, and/or 19 months of age. At each session, infants completed four trials per task (treadmill and walker). We measured step quantity (overall step rate and alternating step rate), step quality (percentage of steps that were alternating, forward, and flat-landing), the ability to self-propel the walker, and caregiver perspectives on both devices.

**Results:** Step quantity increased with age, but infants took more steps in the walker than on the treadmill at all ages. Steps were of equal or higher quality in the walker. By 16 months, half of infants could self-propel. Caregivers viewed both devices favorably, though the majority preferred the walker for home and/or community use.

**Conclusion:** Observational data of overground stepping using a suspension walker suggests overground walkers may be a useful intervention tool for pre-walking infants with Down syndrome.

## Introduction

### Independent walking is delayed in Down syndrome

Down syndrome (DS) is the most common chromosomal condition in the United States and is associated with intellectual disability and significant neuromotor impairments, such as low muscle tone, ligament laxity, reduced muscle strength, and poor postural control.^1,2^ Neuromotor impairments lead to substantial delays in the development of gross motor skills, most notably the development of independent walking. Infants with DS begin walking independently on average around 24 months of age—about one year later than their typically developing peers.^3^ Delays in walking onset restrict children’s participation in their homes and communities and limit learning opportunities that contribute to global development.^4^ Given the importance of independent mobility for participation and development, accelerating walking onset is a priority for families of infants with DS.^5^

The advancement of interventions to improve walking development requires an understanding of the contextual and developmental factors that influence early stepping and walking behavior. This study aims to characterize the development of supported stepping in two contexts—treadmill and overground—to inform the design of intervention approaches for accelerating walking and improving function in infants with DS.

### Observational data informed the development of successful treadmill interventions

In 1986, Esther Thelen demonstrated that seven-month-old typically developing infants could produce alternating steps when held supported over a moving treadmill belt, and later longitudinal work demonstrated that alternating steps became more stable with age.^6,7^ Ulrich and colleagues extended these findings to DS; they found that despite delays in walking development, 11-month-old pre-walking infants with DS could also produce alternating steps when manually supported over a moving treadmill belt, and that alternating steps became more stable with age, as they did in infants with typical development.^8,9^ Together, these observational studies suggested that alternating stepping movements—one of many components of the functional skill of walking—can spontaneously emerge prior to walking onset in a particular physical context. In particular, minimizing balance and postural demands, assisting hip extension via a posteriorly moving ground surface, and providing a tactile stimulus under the feet facilitate stepping behavior, even in infants with DS.

These observational findings led to the development of the most successful known intervention for accelerating walking onset in DS. Researchers capitalized on the stepping behavior elicited by the treadmill and turned it into motor practice. Clinical trials demonstrated that early practice with supported treadmill stepping (supporting infants over a moving treadmill belt for eight min/day, five days/week, starting around 10 months of age) accelerates walking onset by approximately two to four months and improves gait kinematics.^10–15^ Treadmill training protocols leverage neuroplasticity by starting early in development, achieve high total dosage, and provide relatively task-specific practice of stepping movements.

However, current recommendations for pediatric rehabilitation emphasize the importance of active, child-initiated movement and variable practice.^16–21^ On these key ingredients, early stepping training has room for improvement. Treadmill stepping is more passive than real-world walking because stepping movements are elicited purely by the stimulus of the moving belt, with no clear external goals or motivation to move. Moreover, static visual and vestibular information on a treadmill differs from the dynamic sensory input obtained during overground walking; thus, treadmill training fails to provide practice integrating sensory information that is essential for the development of skilled movement.^22–26^

### Mobility devices may support more naturalistic overground stepping practice

Ideally, interventions designed to facilitate walking onset would incorporate properties of real-world overground infant walking. In contrast to the repetitive, isolated stepping produced on a treadmill, natural infant walking is self-initiated and occurs in a variety of physical and social contexts.^27–29^ Contemporary research and theory in typical locomotor development suggests that the variability, distributed practice, and salience that characterize natural overground walking are critical ingredients in the development of functional walking skill.^27–30^

How can properties of natural infant walking be incorporated into early intervention for infants with DS? Mobility devices that provide support for overground stepping, like walkers and gait trainers, provide more immediate access to functional ambulation, in addition to enabling practice of the emergent walking skill. Supported overground walking using gait trainers is commonly implemented by pediatric clinicians to improve walking skill and participation in children with disabilities.^31,32^ However, existing literature primarily focuses on school-age children with cerebral palsy; research on younger populations is scarce^32^ and no studies to date have examined the use of supported stepping devices in infants with DS. It is not known when the ability to functionally walk with a mobility device first emerges. Additionally, prior to the emergence of functional assisted walking, induced overground movement of a supportive mobility device produces a “treadmill-like” stimulus under the feet and may elicit early stepping behavior similarly to treadmill training. To determine whether and at what point in development supported stepping devices are a feasible intervention for infants with DS, it is necessary to understand the developmental trajectory of stepping in this unique context.

### Current study

As reviewed above, treadmill training parameters employed in prior interventions were informed by observational, longitudinal research describing the emergence of treadmill stepping abilities in infants with DS.^8,9^ To assess the potential for overground stepping interventions, we sought to gather analogous foundational data for overground stepping. Thus, we measured stepping behavior longitudinally across two tasks—manually-supported treadmill stepping and harness-supported overground stepping in a suspension walker. To isolate the effects of overground (vs. treadmill) movement, we designed a novel overground paradigm to control for movement speed and direction: We measured stepping as a researcher pushed the walker in a straight path at a prescribed speed. We tested pre-walking infants with Down syndrome across four sessions from 10 to 19 months of age.

Our primary aim was to characterize the developmental trajectory of stepping quantity (overall step rate and alternating step rate) in supported overground stepping trials compared to speed-matched supported treadmill stepping trials across age and motor development. Understanding the emergence of overground stepping is critical to inform the appropriate timing of potential interventions. For example, if overground stepping emerges as early as treadmill stepping, then overground training could start as early as treadmill training. However, if overground stepping emerges later than treadmill stepping or at a particular motor stage, then it may be preferable to start with treadmill training and transition to overground training. To ensure that the walker was not encouraging maladaptive stepping patterns, our secondary aim was to characterize the developmental trajectory of stepping quality (the percentage of alternating steps, the percentage of forward steps, and the percentage of flat-landing steps) across the two tasks. Third, because the walker—unlike a treadmill—allows child-initiated ambulation, we also measured infants’ ability to self-propel in the walker. Our final aim was to assess caregivers’ perspectives on both devices to learn more about whether and when suspension walkers would be accepted by caregivers, and how opinions vary across devices.

## Materials and Methods

### Participants

*N* = 31 pre-walking infants with a diagnosis of DS participated in the study at the University of Southern California (USC) from March 2024 to May 2026 (see table 1 for demographic and medical information). Infants were eligible to participate if they could not yet take 10 walking steps without support and had no additional medical or orthopedic conditions that limited standing or walking. Sessions occurred in one-month windows starting at 10, 13, 16, and/or 19 months of age (e.g., for the 10-month session, infants could be tested between 10 and 11 months). Five infants were born pre-term, so we used adjusted age to determine their session windows. We enrolled infants at the earliest session possible, and invited families to participate in as many sessions as possible. Eight infants participated in one session, 10 infants participated twice, 9 infants participated in three sessions, and 4 infants participated in all four sessions, for a total of *N* = 71 sessions (15 10-month, 21 13-month, 17 16-month, 18 19-month; see supplemental figure 1). One additional infant enrolled but is not included because they did not complete the 10-month session and did not return for another visit; we also excluded one 13-month session from one of the 31 included participants because the infant was too fussy to complete the session.

**Table 1.**
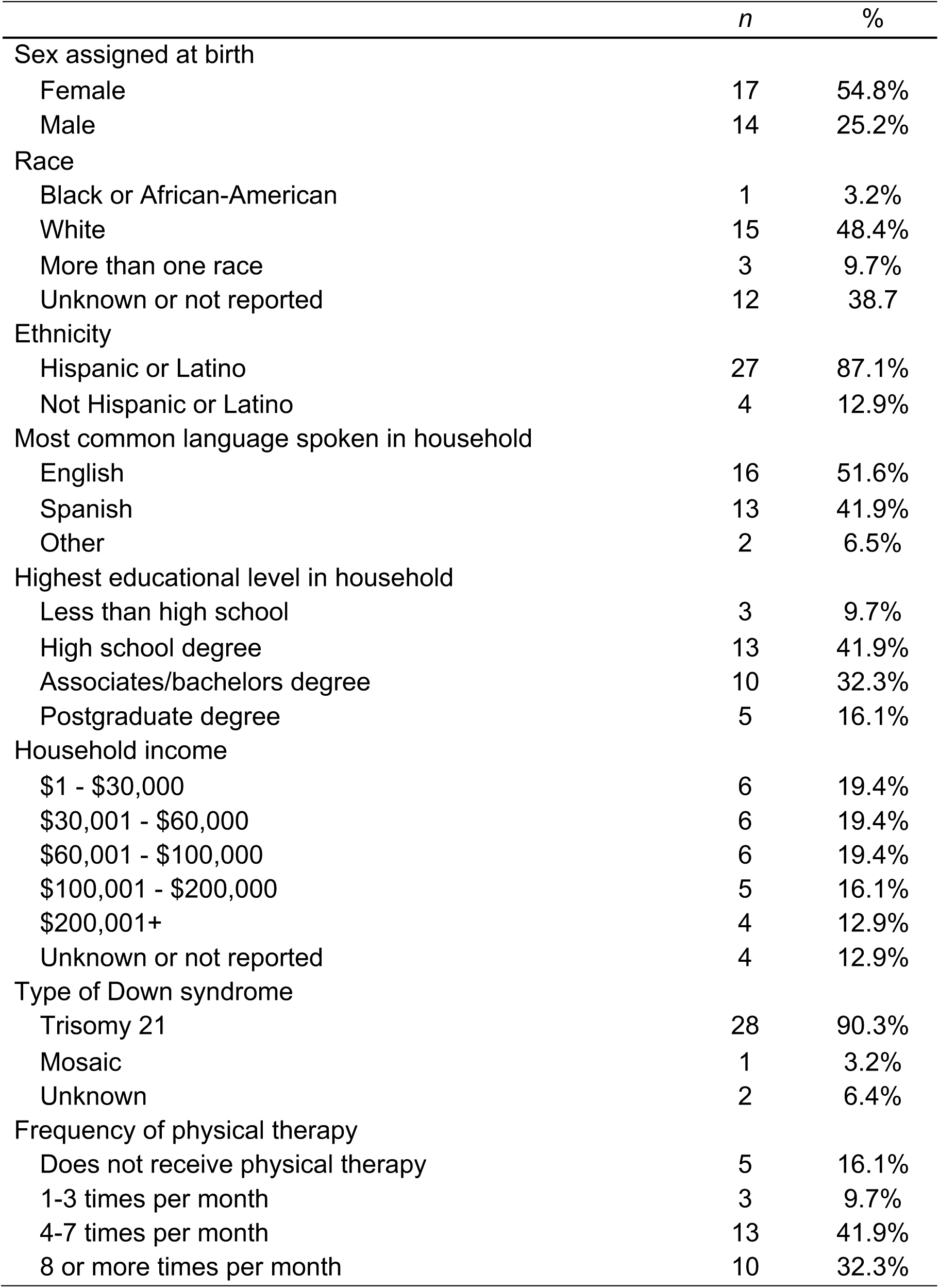
Demographic and medical information. All responses were recorded at the infant’s first session.

The procedure was approved by the USC IRB (UP-22-01092). Caregivers provided informed consent at the first session and were able to withdraw or skip future sessions at any time. Families received a $25 Amazon gift card per session and were provided with free parking or transportation reimbursement. We recruited infants primarily through medical records from Children’s Hospital Los Angeles. Study data were collected and managed using REDCap electronic data capture tools hosted at USC.^33,34^ The funder played no role in the design, conduct, or reporting of this study. Processed data, data collection and behavioral coding protocols, and analysis code are shared on Databrary at [URL TO BE ADDED], as is video data of each session (according to caregiver permission).

## Materials and procedure

Sessions lasted approximately 90 minutes. Sessions were conducted in English or Spanish per caregiver preference. At each session, the infant completed: (1) *four treadmill trials*: two slow (0.1 m/s) and two fast (0.2 m/s) stepping trials; (2) *four overground trials*: two slow (0.1 m/s) and two fast (0.2 m/s) stepping trials; (3) *one overground propulsion test*; and (4) a *gross motor function assessment*. During the gross motor function assessment, caregivers filled out questionnaires to provide demographic and medical history and report their perspectives on both devices. Infants were also recorded in five minutes of free play with their caregiver with and without the walker; these data are reported separately.^35^ Trial speeds were selected to mirror those of prior treadmill protocols.^8,9^ We counterbalanced task order and speed, for a total of four possible condition orders. The assessment of gross motor function was always performed last to minimize fatigue during the stepping tasks. Infants wore lab-provided soft shoes with rubber soles during the stepping tasks to protect the toes from injury when dragging on the floor or treadmill; shoes were removed for the gross motor function assessment.

### Treadmill trials

We replicated the treadmill procedure outlined in Ulrich et al.^8,9^ Infants were suspended in an upright position over a GaitKeeper Mini treadmill (LiteGait®; 24” wide x 47” long x 6” high, 50 lbs). Infants were supported manually at the axillae by the researcher unless the infant was too fussy to be held by a stranger, in which case the infant was held by their caregiver; infants were held such that the infant supported their own weight on their feet as much as possible (figure 1A). The treadmill speed was set to 0.1 m/s or 0.2 m/s using an attached remote, and trials lasted 30s each. The treadmill trials were video recorded from three stationary views (frontal, posterior, and diagonal) that were synced online using OBS software.

**Figure 1.** Exemplar pictures of the two tasks. (A) Treadmill: Infant is held supported at the axillae as the treadmill belt moves at 0.1 m/s or 0.2 m/s. Infants complete two trials at each speed. (B) Walker: Infant wears an adjustable harness that clips into the overhead suspension bar. The researcher pushes the walker at 0.1 m/s or 0.2 m/s, to match the speed of the treadmill belt. Infants complete two trials at each speed.

### Overground trials

We used a Kaye suspension walker, a wheeled posterior walker with a mounted overhead bar (KAYE Products, Inc., 42” high, 24” x 23” footprint; figure 1B). The researcher outfitted the baby into an adjustable soft harness, and then clipped the straps of the harness onto the overhead bar. The researcher adjusted the length of the harness straps so that the infant could maintain a fully upright position while bearing weight through the feet with knees and hips extended (figure 1B).

The overground stepping trials were performed in an indoor hallway (1.2m wide x 9m long). The researcher pushed the walker at 0.1 m/s and 0.2 m/s by traveling 3m and 6m in 30s, respectively. To maintain the correct speed, the ground was taped in 1.5m increments and the researcher carried a timer in hand (e.g., the researcher verified they were crossing each 1.5m mark in ∼7.5 seconds for the 0.2 m/s trials). For the overground stepping trials, the front wheels of the walker were locked in a straight-ahead position to facilitate straight-path pushing. The stepping trials were video recorded from a moving frontal view (assistant with an iPad on rolling dolly).

### Overground propulsion test

After the four overground stepping trials were complete, we assessed the infant’s ability to self-propel in the walker. For propulsion trials, the front wheels of the walker were locked in a straight-ahead position to minimize steering demands and the back wheels’ posterior roll stops were engaged to prevent backwards movement. The researcher placed the walker at the first line of tape on the ground and asked caregivers to encourage the infant to walk toward them, using vocal cues and toys as incentives. We recorded whether the infant was able to propel the walker or not. No assistance was provided to move the walker. We ended propulsion trials after the infant propelled 3m or after 60s elapsed, whichever came first. The propulsion trials were video recorded from a diagonal frontal view (assistant with a handheld iPad).

### Gross motor function assessment

Because gross motor function varied within and across age groups, we administered the Gross Motor Function Measure-88 (GMFM-88), a criterion-referenced assessment of gross motor function that is valid and reliable (*ICC* = 0.95) in children with DS.^36^ Specifically, we administered three of the five dimensions: B. Sitting, D. Standing, and E. Walking/running/jumping. Each item is rated on a 4-point scale, and scores for each dimension are obtained by summing item scores. We did not administer the “Lying & rolling” or “Crawling & kneeling” dimensions due to time constraints. Testing took approximately 15-20 minutes in total. The GMFM was scored live by the primary assessor, and a second assessor scored one random session from each participant. Reliability was high for each dimension (*ICCs* > .94).

### Caregiver perspectives

At the end of each session, caregivers completed a custom “Equipment Questionnaire” to understand their perspective on both devices; the survey was added after data collection started, so data from six sessions is missing (see colored key in supplemental figure 1 for specifics on which sessions did not have caregiver perspectives data). The questionnaire consisted of six Likert-scale questions (three per device) about their child’s enjoyment, their interest in using the device in their home or community, and how easy they thought it was for their child to use. We also asked two optional open-ended questions about what they liked and/or disliked about the walker, including what features they found helpful and what features could be added or improved. For the latter half of sessions (32 sessions from 17 infants), we added a forced-choice question asking which device (treadmill or walker) they would choose to have for home use and an optional open-ended question about why they selected that device.

### Data coding and processing

Coders first annotated the start and end of each trial using Datavyu video-coding software (datavyu.org). We calculated the true speed of the walker trials by dividing the distance traveled by the trial time.

Next, coders annotated stepping activity. For each of the stepping trials, coders scored each step by identifying the moment each foot lifted off the ground/treadmill and regained contact with the ground/treadmill. Left and right steps were coded separately. A primary coder annotated all four trials per task and a second coder independently annotated at least one trial per task to test inter-observer reliability. We classified individual steps as alternating if the onset of a step on one foot occurred at the same time or later than the offset of the previous step on the other foot. A primary coder annotated all trials per task and a second coder independently annotated at least one trial per task to test inter-observer reliability. Disagreements were resolved by discussion. Reliability on number of steps per trial (*ICC* = 0.995) and number of alternating steps per trial was high (*ICC* = 0.993). We removed 323 steps that were longer than 2s (out of 11,297 coded steps) because these represented times when infants held their feet off the ground rather than stepped. We calculated the *overall step rate* (steps/min) and *alternating step rate* (alternating steps/min) by counting the total number of left and right steps in each trial and dividing by the trial time.

As a metric of stepping quality, we also calculated the *percentage of alternating steps* by dividing the number of alternating steps by the total number of steps (we report alternation as both a rate and a percentage because they provide different information; for example, an infant could have a low alternating step rate because they did not take many steps overall, but still have a high percentage of alternating steps). The other two metrics of stepping quality were calculated from a second coding pass, which was completed after coders met to review and resolve step disagreements. In this pass, coders scored two dichotomous variables per step—how the step moved through space and how it landed. Steps were marked as “forward” if the step moved forward through space or lifted and landed in place (steps not counted as forward included steps where the infant tapped the foot in front of the body, dragged the foot behind the body, took a backwards step, or moved the foot through the air in multiple directions). Steps were marked as “flat” if the foot landed flat within one second of initial contact (steps not counted as flat included steps that landed on the toes, heel, side of the foot, or opposite foot). A primary coder annotated all steps per task and a second coder independently annotated 25% of the steps per task (every fourth step). Disagreements were resolved by discussion. Reliability was high for type (κ = 0.88) and landing (κ = 0.94). We calculated the *percentage of forward steps* and the *percentage of flat steps*.

## Data analysis

An a priori power analysis suggested a sample size of 20 infants was sufficient to detect moderate statistical associations.

We ran separate statistical models for all five stepping outcomes (overall step rate, alternating step rate, percentage of alternating steps, percentage of forward steps, percentage of flat steps). For step rate models, we fit a linear mixed model with trial-level data using the *lme4* package (Version 2.0.1) in R. For percentage models, we fit generalized linear mixed models with a beta-binomial distribution (to account for percentage data exhibiting overdispersion) and logit link using the *glmmTMB* package (Version 1.1.14) in R; fixed effect coefficients were exponentiated to obtain odds ratios (*OR*s) for ease of interpretation.

Occasionally, extra trials were administered due to experimenter error (2 instances on the treadmill, 9 instances in the walker). In these cases, to capture infants’ best performance^27,37,38^, we included the two trials with the highest alternating step rate. (As a sensitivity analysis, we re-ran models including all valid trials; this did not change the pattern of results.) Additionally, although we attempted to match the speeds as precisely as possible between conditions, the actual speeds in the walker condition were occasionally incorrect due to experimenter error or because the infant self-propelled faster than the intended speed. Because this could have inflated the stepping outcomes in the walker condition, we excluded 34 walker trials where the speed was ≥0.025 m/s different from the intended speed (33 trials faster than intended, 1 trial slower than intended). Thus, we modeled data from a total of 543 trials.

Across models, fixed effects included infant age in months, task (treadmill = 0, walker = 1), and speed (slow = 0, fast = 1) as well as the age×task interaction. Because models included interaction effects, all predictors were centered at the mean so that coefficients represent main effects (*M*_age_ = 15.2 months). To account for individual differences in baseline stepping and developmental trajectories, random effects included a random intercept and random age slope for participants and a random intercept for sessions nested within participants. For the alternating step rate model, estimation of the correlation between the participant-specific intercept and age slope resulted in a singular fit; therefore, these random effects were specified as uncorrelated for the alternating step rate model. For significant interactions, we used the *emmeans* package (Version 2.0.3) in R to test the effect of condition at each of the session ages (10, 13, 16, and 19 months).

Due to large inter-individual differences in motor skill development in DS, we also tested the effects of gross motor function, rather than chronological age, on our primary outcome of stepping quantity (overall step rate and alternating step rate). We repeated the analyses substituting each dimension of the GMFM (sitting, standing, and walking/running/jumping) for age in three separate models per outcome variable, but we included only a random intercept for participants and a random intercept for sessions nested within participants (we removed the random age slope due to singularity and convergence issues with these GMFM models). Models were fit using maximum likelihood and used the same observations. We compared the fit of each GMFM model with the corresponding age model using the Akaike Information Criterion (AIC). Lower AIC values were interpreted as better relative fit among candidate models, accounting for both model complexity and goodness of fit.^39,40^

The binary ability to self-propel was analyzed using a generalized linear mixed model (binomial distribution with logit link) using the *lme4* package in R. We tested age (mean-centered) as a fixed effect and included a random intercept for participant. As with step quantity outcomes, we repeated the analysis substituting each dimension of the GMFM (sitting, standing, and walking/running/jumping) for age in three separate models.

For the caregiver perspective data, Likert responses were dichotomized for all six survey questions (agree/strongly agree vs all other responses, and somewhat/very easy vs somewhat/very difficult). Because questions were repeated across devices, we ran one model for each question type (child enjoyment, ease of use, interest in home/community use). We fit generalized linear mixed models (binomial distribution with logit link) using the *lme4* package in R. We tested age (mean-centered), device (dummy-coded and mean-centered, treadmill = 0, walker = 1), and the age×device interaction as fixed effects and included a random intercept for participant. Open-ended responses were not formally analyzed, but examples and informal summaries are presented to add context to the quantitative responses.

## Results

### Developmental trajectory of stepping quantity across tasks

#### Overall step rate

The intercept—representing the step rate at the average age, task, and speed—was 32 steps/min (figure 2A, table 2). Step rate increased with age and speed and was higher in the walker than the treadmill. At the average age and speed, infants took 18 additional steps per minute in the walker compared to the treadmill. Moreover, the age×task interaction was significant; step rates were higher in the walker at all age points (*p*s < .001), but the effect was larger at older ages. Of note, infants took zero steps in only 22 of the 543 trials (8 treadmill trials, 14 walker trials).

**Figure 2.**
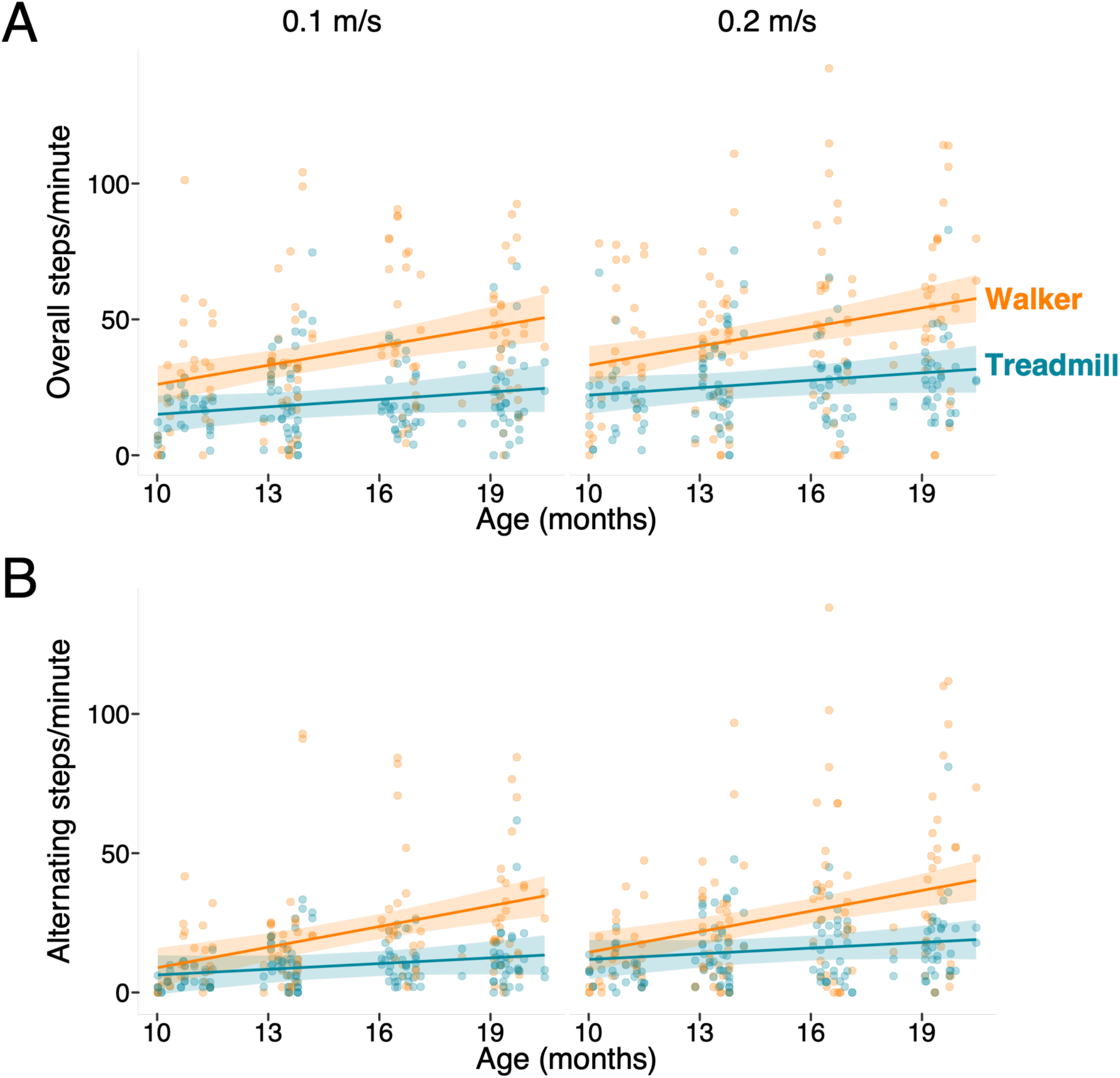
Developmental trajectory of step quantity: (A) Overall step rate and (B) alternating step rate as a function of age (x-axis) and task (colors; blue = treadmill, orange = walker). Banded lines represent the model estimation with 95% confidence interval. Circular points represent trial-level individual data from each infant (repeated observations clustered within sessions and infants). Across (A) and (B), the left panel includes data from the slower trials (0.1 m/s) and the right panel includes data from the faster trials (0.2 m/s).

**Table 2.**
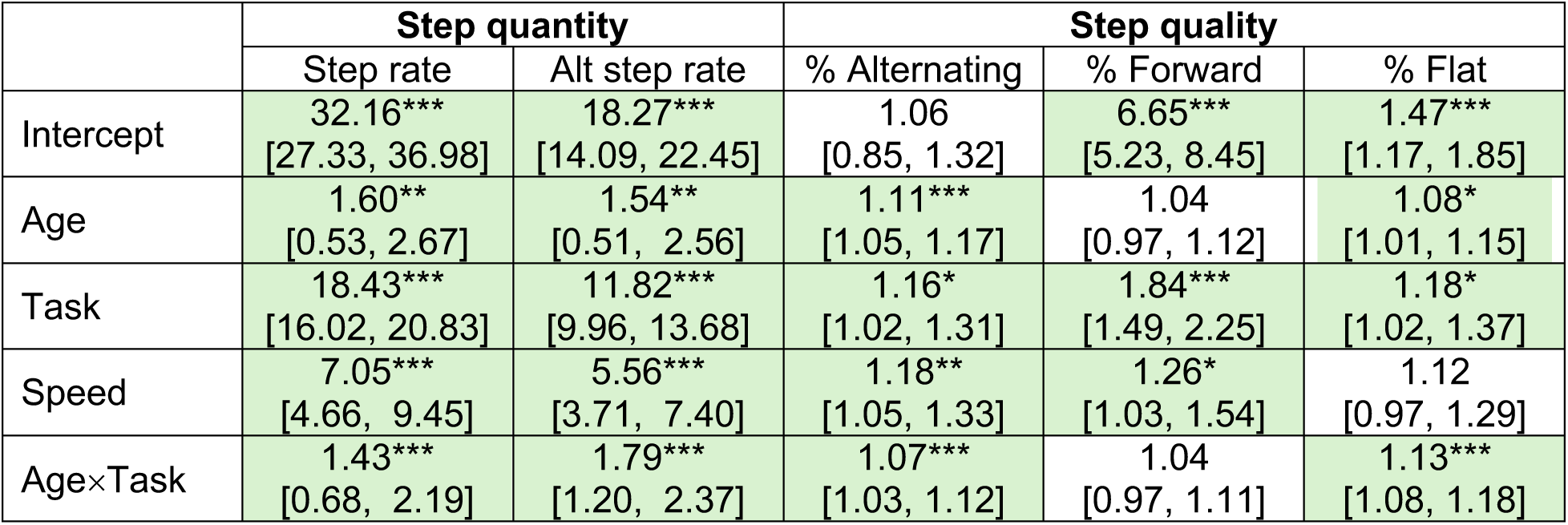
Summary of model results for step quantity and step quality. For step quantity (overall step rate and alternating step rate), values listed are coefficient estimates with the upper and lower bounds of the 95% confidence interval in parentheses. For step quality (percentage of alternating steps, percentage of forward steps, and percentage of flat landing steps), values listed are odds ratios with the upper and lower bounds of the 95% confidence interval in parentheses. Age (months) was centered at the mean. Task was dummy-coded (treadmill = 0, walker = 1) and centered at the mean. Speed was dummy-coded (slow = 0, fast = 1) and centered at the mean. A random intercept and random age slope was included in all models. Cells are highlighted in green if significant. Asterisks denote *p*-values: \**p* < .05, \*\**p* < .01, \*\*\**p* < .001.

### Alternating step rate

As in prior treadmill studies, infants demonstrated a variety of step patterns, including repeated steps on the same foot and simultaneous steps on both feet. Therefore, the alternating step rate was lower than the overall step rate. The intercept—representing the alternating step rate at the average age, task, and speed—was 18 steps/min (figure 2B, table 2). Nonetheless, overall rate and alternating step rate were highly correlated (*r* = .87), so the models followed a similar pattern: Alternating step rate increased with age, speed, and was higher in the walker than the treadmill. At the average age and speed, infants took 12 additional alternating steps per minute in the walker compared to the treadmill. Moreover, the age×task interaction was significant; alternating step rates were higher in the walker at 13 months and beyond (*p*s < .001), and the task effect was larger at older ages.

### Effects of gross motor function on stepping quantity

#### Overall step rate

Both the standing and walking dimension models followed the same pattern as the age model (supplemental figure 2), but there was no significant dimension×task interaction (*p* = .57) in the sitting model. Of all four models, the walking dimension model had the lowest AIC (4565), indicating the best relative fit, compared to models containing standing skill (ΔAIC = 18), age (ΔAIC = 20), or sitting skill (ΔAIC = 37).

### Alternating step rate

All three GMFM dimension models followed the same pattern as age for alternating step rate (positive main effects and significant development×task interactions; supplemental figure 3). Like overall step rate, the walking dimension model also had the lowest AIC (4259) for alternating step rate, indicating the best relative fit compared to models containing standing skill (ΔAIC = 41), age (ΔAIC = 58), or sitting skill (ΔAIC = 87).

## Developmental trajectory of stepping quality across tasks

For the following three stepping quality models, data was missing from the 22 trials where infants took zero steps (8 treadmill trials, 14 walker trials), so we effectively modeled data from 508 trials for the following outcomes.

### Percentage of alternating steps

At the average age, task, and speed, the probability of an alternating step was 52% (figure 3A, intercept from table 2 transformed with inverse-logit function). The odds of an alternating step was higher at older ages, and at faster speeds. The task effect varied by age (main effect of task and age×task interaction): At 16 and 19 months, steps were more likely to be alternating in the walker than the treadmill (*OR*s of 1.23 and 1.52, respectively; *p*s < .002), but there was no statistically significant difference in the likelihood of an alternating step between tasks at 10 (*p* = .08) and 13 months (*p* = .91).

**Figure 3.**
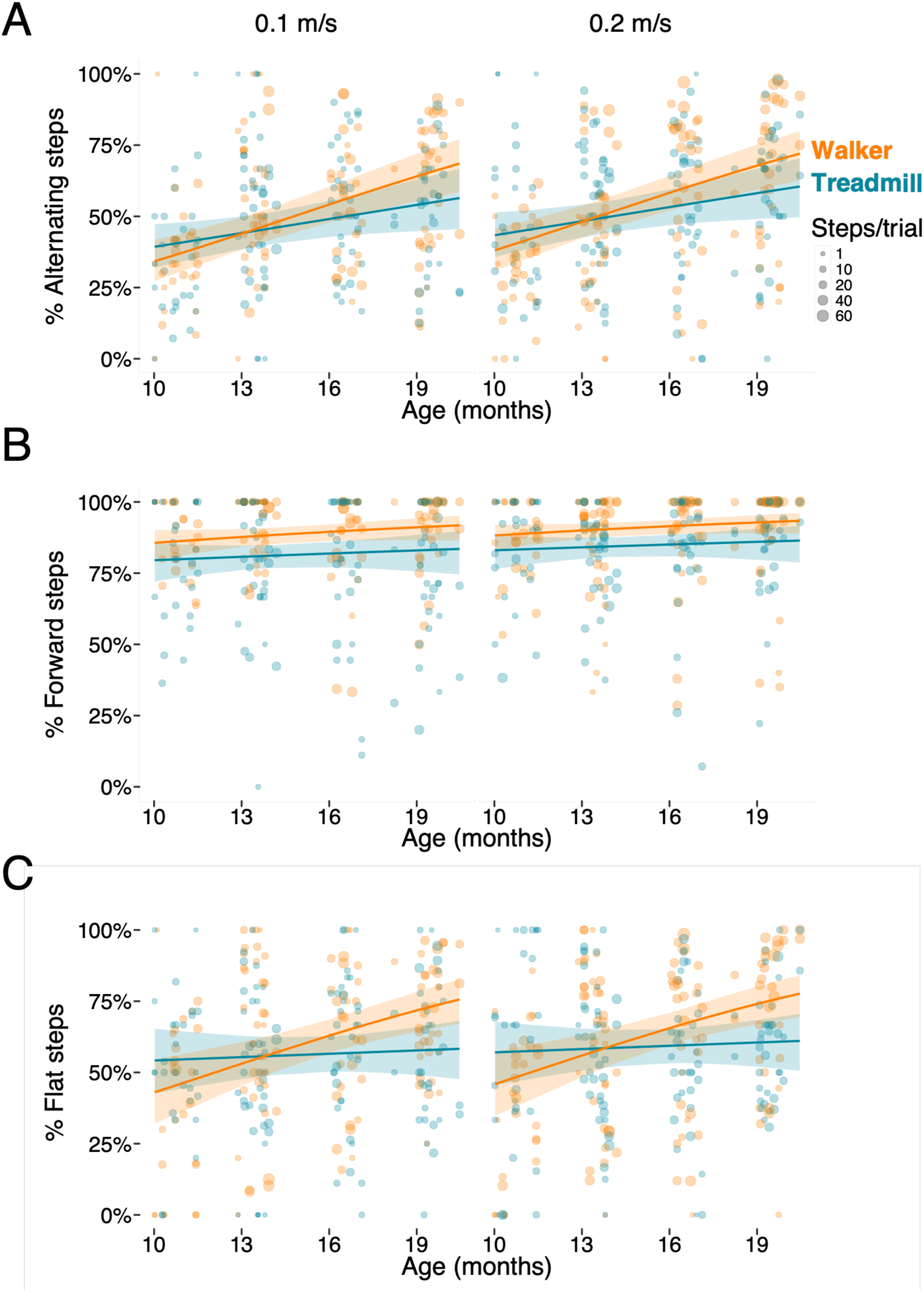
Developmental trajectory of step quality: (A) Percentage of alternating steps, (B) percentage of forward steps, and (C) percentage of flat-landing steps as a function of age (x-axis) and task (colors; blue = treadmill, orange = walker). Banded lines represent the model estimation with 95% confidence interval. Circular points represent trial-level individual averages from each infant; the size of circular points represent the number of steps used to calculate the average for that trial. Across (A), (B), and (C), the left panel includes data from the slower trials (0.1 m/s) and the right panel includes data from the faster trials (0.2 m/s).

### Percentage of forward steps

At the average age, task, and speed, the probability of a forward step was 87% (figure 3B, intercept from table 2 transformed with inverse-logit function). Despite high rates of forward steps generally, the likelihood of a forward step was still higher in the walker compared to the treadmill, and at faster speeds compared to slower speeds. There was no main effect of age or age×task interaction.

### Percentage of flat steps

At the average age, task, and speed, the probability of a flat-landing step was 70% (figure 3C, intercept from table 2 transformed with inverse-logit function). At 16 (*OR* = 1.39, *p* < .001) and 19 months (*OR* = 1.76, *p* < .001), steps were more likely to land flat on the walker than treadmill; there was no difference at 10 (*p* = .11) or 13 months (*p* = .15).

## Self-propulsion in the walker

The ability to self-propel also improved over time. At 10 months, none of the 15 infants were able to self-propel. At 13 months, 6/21 infants (29%) self-propelled; at 16 months, 7/17 infants (41%) self-propelled; and at 19 months, 10/18 infants (56%) self-propelled. The linear mixed model confirmed that the odds of self-propulsion increased significantly with age (*OR* = 1.67, *p* = .01). As with step quantity, we tested the effects of gross motor function (rather than age) on propulsion by repeating our model substituting each dimension of the GMFM (sitting, standing, and walking/running/jumping) for the age predictor. All models estimated positive associations between developmental status and the ability to self-propel, but the main effect of the standing dimension was not significant (*p* = .07). Of all four models, the sitting dimension model had the lowest AIC (70), indicating the best relative fit compared to models that contained age (ΔAIC = 6), walking skill (ΔAIC = 8), or standing skill (ΔAIC = 13).

## Caregiver perspectives

Caregivers’ perspectives of both devices were largely positive (figure 4). Collapsed across ages, most caregivers agreed that their child enjoyed the treadmill (63%) and walker (60%), and that the treadmill (54%) and walker (59%) were easy for their child to use. Caregivers also reported that they would be interested in using a treadmill (69%) and walker (82%) in their home or community. Results from the linear mixed models indicate that there were no significant differences in response by age, between devices, or for the age×device interaction, across all three questions.

**Figure 4.**
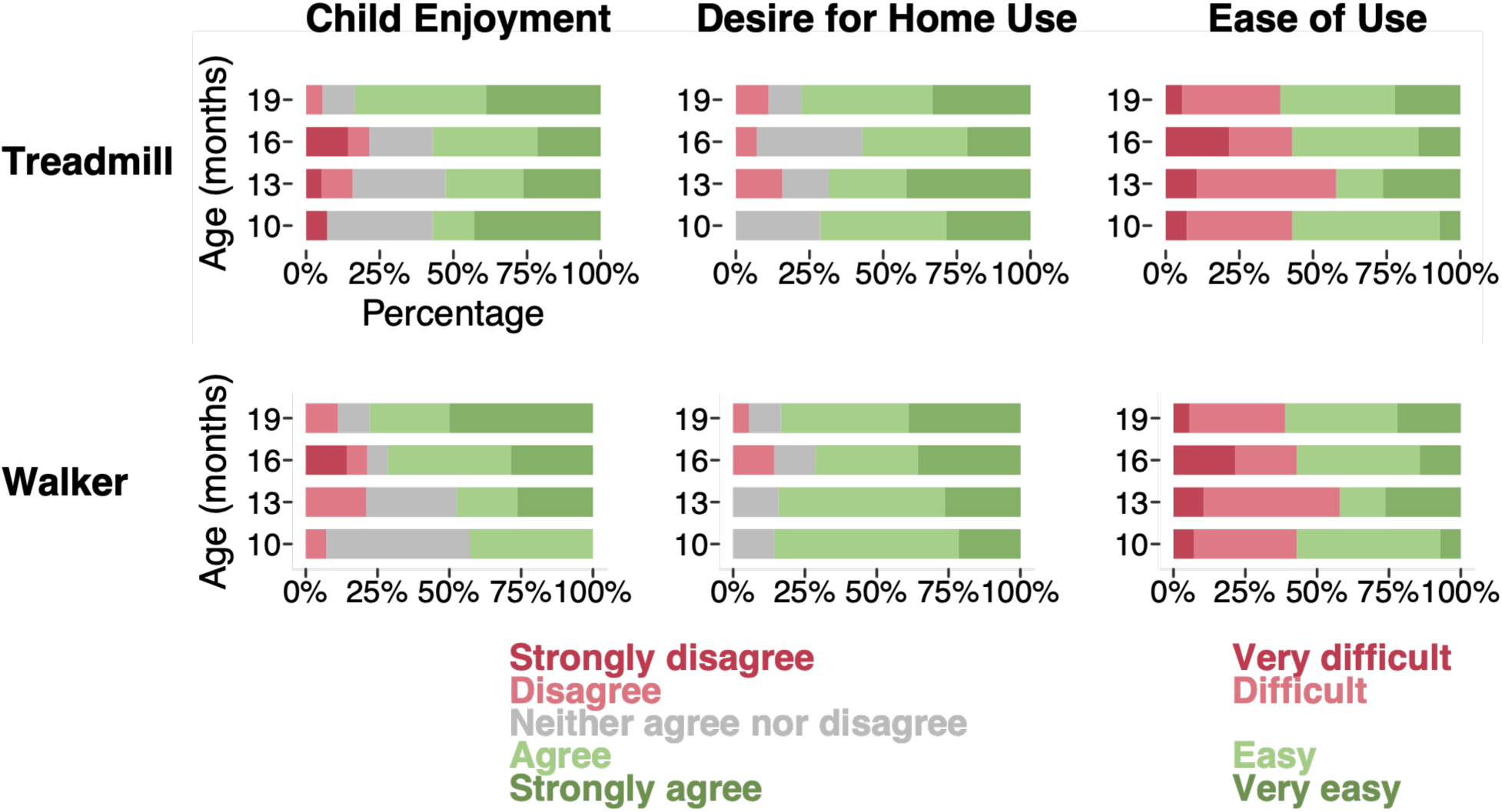
Caregiver perspectives on the treadmill (top row) and walker (bottom row). The first column illustrates caregiver responses regarding child enjoyment (“My child enjoyed using the [treadmill/walker].”). The second column illustrates caregiver responses regarding home use (“I would be interested in using a [treadmill/walker] in my own home or community.”). The last column illustrates caregiver responses regarding ease of use (“How easy do you think it was for your child to use the [treadmill/walker]?”).

In open-ended responses regarding what they liked about the walker, caregivers highlighted how the walker provided their child with a new perspective (e.g., *“He could see higher and reach higher objects*”) and independence (e.g., *“My son seems interested and happy to walk without help and can discover different spaces independently.”*). Many parents also indicated that it was easier for their child to step (e.g., *“They could stand and give a few steps. They have never done that before.”*). When asked what they disliked about the device, caregivers tended to reference the physical footprint of the device (e.g., *“It should be a little smaller in case they have small spaces at home.”*) and features of the harness itself (e.g., *“Complex buckles means baby could get frustrated before they begin.”*)

For the subset of sessions in which caregivers were asked to select which device they would choose to have for home use, caregivers selected the walker 78% of the time. In the open-ended response, most caregivers indicated that they chose the walker because it was more practical, either due to space constraints (e.g*., “We don’t have space for a treadmill but could use the walker at the park.”*) or the physical design (e.g*., “The suspension walker seems easier since she is supported by the harness and not by parents’ arms.”*).

## Discussion

In sum, infants demonstrated readiness for overground stepping as early as treadmill stepping. Infants took more steps in the walker compared to the treadmill at all ages, the walker advantage grew over development, and the effect was large: Infants produced over 50% more steps per minute in the walker compared to the treadmill. Moreover, the effect of the walker was consistent across outcome measures; at all ages, steps were of equal or higher quality in the walker compared to the treadmill, so the walker did not encourage maladaptive stepping patterns. Infants were able to self-propel the walker (a feature not possible on the treadmill) as early as 13 months of age, and caregivers rated the walker positively. Together, results suggest overground walkers demonstrate high potential for early intervention for pre-walking infants with Down syndrome.

## Potential mechanisms of improved stepping in the walker

Why did infants step more in the walker? Although our design does not allow us to determine why infants stepped more in the walker than on the treadmill, we can hypothesize a few mechanisms. Based on prior work with typically developing infants, there is evidence that optic flow consistent with self-motion increases infant stepping.^41,42^ Thus, it is plausible that the sensory experience provided by the walker (i.e., optic flow plus vestibular stimulation consistent with movement) increases stepping relative to the mismatched experience provided by the treadmill. Moreover, by physically moving through space rather than remaining in one place on the treadmill, the experience of moving in the walker may be more interesting or motivating than being supported over the treadmill, leading to increased arousal and engagement; increased arousal is also associated with increased stepping and kicking in typically developing infants.^43^ Finally, the postural support provided by the harness may be more comfortable or supportive than being held supported under the axillae, as is done on the treadmill, which could have affected infants’ responses.

Infants’ step quality was also of equal or higher quality in the walker compared to the treadmill. Forward stepping showed the strongest task effect, but no interaction with age, whereas alternating and flat stepping showed smaller task effects that increased with age. The forward stepping effect may therefore reflect implicit task differences that do not change as infants develop; for example, the sensory experience of forward progression provided by the walker (which the treadmill does not provide because infants remain stationary) may encourage more forward-directed leg movements. In contrast, the age-related increases in alternating and flat-footed stepping may reflect infants’ growing ability to capitalize on the support and opportunities provided by the walker. Alternation and controlled foot placement require coordination of postural control, weight transfer, and limb movements; these capacities improve with motor development, but may also be easier to elicit with the physical support of the walker versus the treadmill (i.e., distributed harness support versus manual support under the axillae).

Future work should directly explore these possible explanations for differences in both stepping quantity and quality to elucidate which factors act as true mechanisms of improved stepping in the walker.

## Application to clinical practice and intervention

Our current design has several strengths and provides useful foundational data for designing future interventions. The within-session comparison of treadmill and walker stepping reduces between-child confounding. Moreover, although we restricted the potential opportunities of the walker by having a researcher push the walker in a straight path at a prescribed speed, these controlled trials allow for fair, direct comparison between tasks. Last, the partially longitudinal design spans clinically relevant ages and captures age-related changes in stepping behavior.

Not only did the walkers encourage more and often higher quality stepping than the treadmill, but the practice provided by the walker is also more task-specific to real-world walking. Specificity is defined as the “extent to which practice conditions mirror the sensorimotor, contextual, physiological, cognitive, and psychological demands of the target behavior so that the neurobehavioral processes elicited during practice facilitate effective and persistent real-world walking.”^44^ Because infants move through physical space in the walker, the walker may better promote the perceptual-motor processes needed for overground walking compared to treadmill stepping. In less controlled settings where the infant could control walker movement (allowing them to turn, start, and stop), the walker would provide even more task-specific practice, and our results on propulsion suggest infants would be able to self-propel the walker as early as 13 months of age. Additionally, caregiver preference for the walker in the home and/or community highlights that the walker may also be more practical for everyday life.

We found that measures of motor skill (as measured by the GMFM) were a better predictor of infants’ stepping abilities than their age, aligning with prior work demonstrating that infants’ capacity, skill, and/or experience is a better predictor of infants’ behavior than age itself.^45–47^ Interestingly, for stepping quantity, models including walking skill (Dimension E of the GMFM) demonstrated the best fit for overall and alternating step rate, whereas for propulsion, the model including sitting skill (Dimension B of the GMFM) was the best fit. The difference in results may suggest that self-propulsion depends largely on trunk stability so that infants can generate the necessary force to move the walker, whereas stepping quantity benefits more from advanced upright locomotor abilities and leg strength. However, regardless of whether we used age or skill to predict infants’ stepping quantity, infants stepped more in the walker than on the treadmill. Thus, there is no reason to wait until certain clinical criteria are met to start overground training—infants are ready for overground stepping as early as 10 months, just as they are ready for treadmill training in our and prior work.

Finally, a common concern for commercial baby walkers is that they may encourage toe-walking or other maladaptive walking patterns. Although we used a medical-grade walker that allows for proper anatomical positioning, similar concerns may still exist for parents or clinicians when deciding whether to introduce an overground mobility device. However, our results on stepping quality—that steps were equally or more likely to be alternating, move forward, and land flat in the walker compared to the treadmill—suggest that maladaptive walking patterns are not a concern, at least for the medical-grade walker that we used.

## Limitations and future directions

This was an observational study testing immediate stepping behavior, so it cannot determine whether overground stepping accelerates walking onset as has been found in prior treadmill stepping work. Because we found that infants stepped more in the walker than on the treadmill when measured in brief laboratory trials, we hypothesize that prolonged practice with the overground walker may accelerate walking onset as much as, or potentially more than, treadmill training. Findings from the current study will inform the design of future home-based interventions to directly test the effects of prolonged stepping practice in the overground walker on walking development, as well as the feasibility of home and community use.

Another limitation was that longitudinal follow-up was unbalanced and caregiver perspective data was missing for some visits. We enrolled infants at the earliest session possible but initial session(s) are missing if infants enrolled after 10 months. Later sessions are missing if families were too busy to return for additional sessions, if we lost contact with the families, or if infants began to walk independently before 19-months. Moreover, caregiver perspective data was missing for some visits, so current findings should be treated as preliminary.

Despite the robustness of the task effect on stepping behaviors, infants nonetheless displayed considerable interindividual variability. In future work, we plan to explore these individual differences to learn more about what factors predict various response patterns to support a precision rehabilitation approach.

## Conclusions

Our overarching goal in conducting the current study was to gather foundational observational data on the development of overground stepping in a suspension walker akin to that used to develop effective treadmill training parameters. Our findings—that pre-walking infants with Down syndrome step even more in overground walkers than they do supported on a treadmill—present a compelling case for the potential of overground suspension walkers and provide the data needed to develop informed overground stepping interventions in the future.

## Acknowledgments

This project was supported by a Pediatric Research Grant from the Foundation for Physical Therapy Research. We are incredibly grateful to the children and families who participated in this research. We thank Lauren Bussell, Melanie Paiz, Marcus Partida, Valeria Reyes Bastidas, and Lauren Vasquez for their contributions to data collection and coding.

## Declaration of interest

The authors report that there are no competing interests to declare.

## Data availability statement

Processed data, protocols, and analysis code are shared openly on Databrary at [URL TO BE ADDED]. Video data of each session is shared according to caregiver permission on Databrary at [URL TO BE ADDED].

## Figures

Figure removed for preprint. Please contact the corresponding author for an image of the treadmill and suspension walker.

## Notes

### Competing Interest Statement

The authors have declared no competing interest.

### Author Declarations

The IRB of the University of Southern California gave ethical approval for this work.

### Summary of Updates

Changes have been incorporated into the introduction and discussion based on a peer reviewer's comments. Statistical analyses have been updated and described in more detail.

