## Supplement for "Overground suspension walkers elicit more and higher quality stepping than treadmills: A longitudinal study of pre-walking infants with Down syndrome"

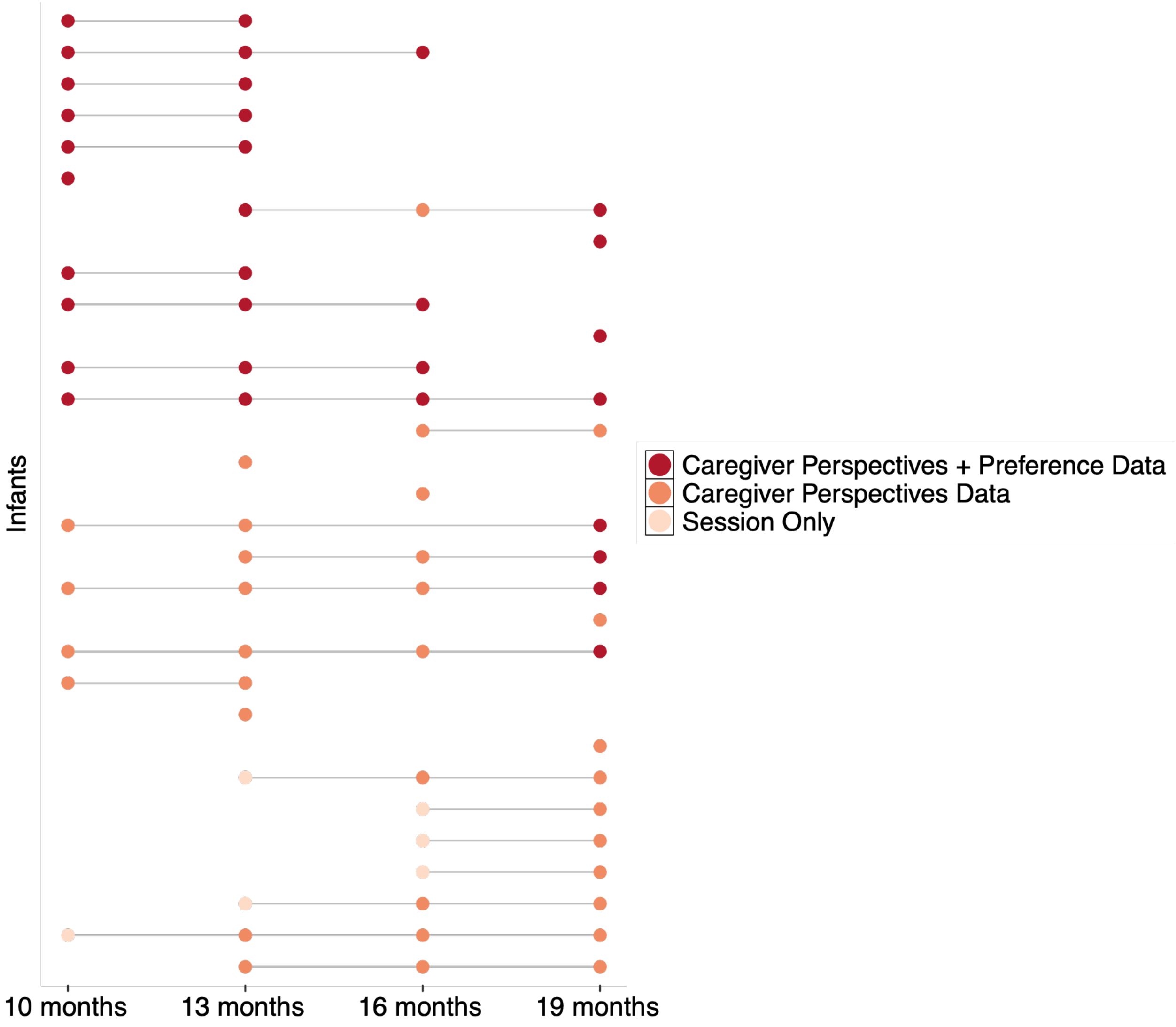


**Supplemental Figure 1**. Sessions completed by each participant, indicated by a filled circle. Colors indicate whether the session included no caregiver perspective data (light peach), had perspective data but no question on device preference (orange), or had both perspective data and device preference (red). Infants are ordered by enrollment (earliest enrollment at bottom).

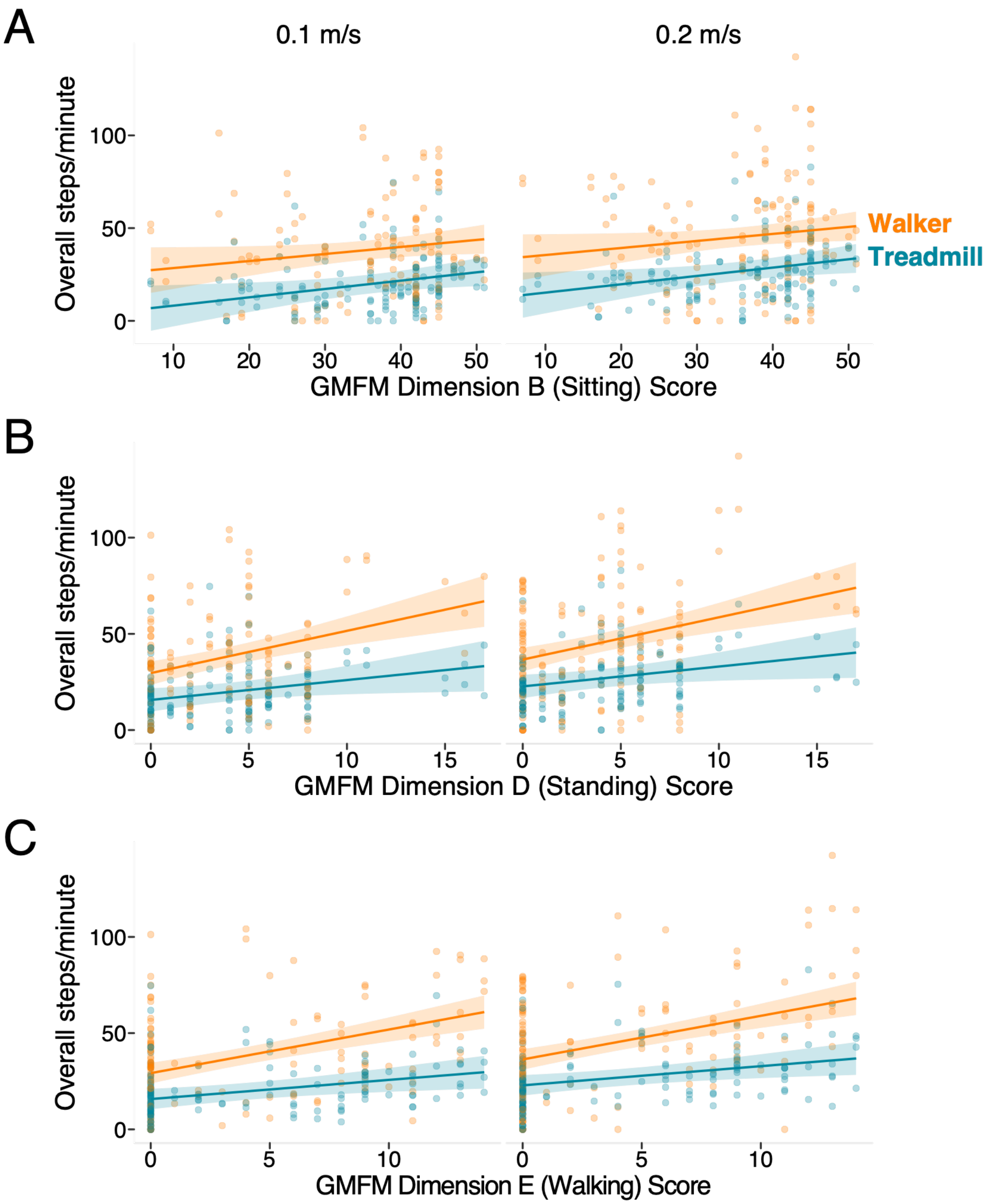


**Supplemental Figure 2**. Developmental trajectory of overall step rate as a function of gross motor skill (x-axis) and task (colors; blue = treadmill, orange = walker): (A) Sitting skill (Dimension B of the GMFM); (B) standing skill (Dimension D of the GMFM); (C) Walking skill (Dimension E of the GMFM). Banded lines represent the model estimation with 95% confidence interval. Circular points represent trial-level individual data from each infant. Across (A), (B), and (C), the left panel includes data from the slower trials (0.1 m/s) and the right panel includes data from the faster trials (0.2 m/s). Models follow the same overall pattern as the age model reported in paper (Figure 2A and Table 2).

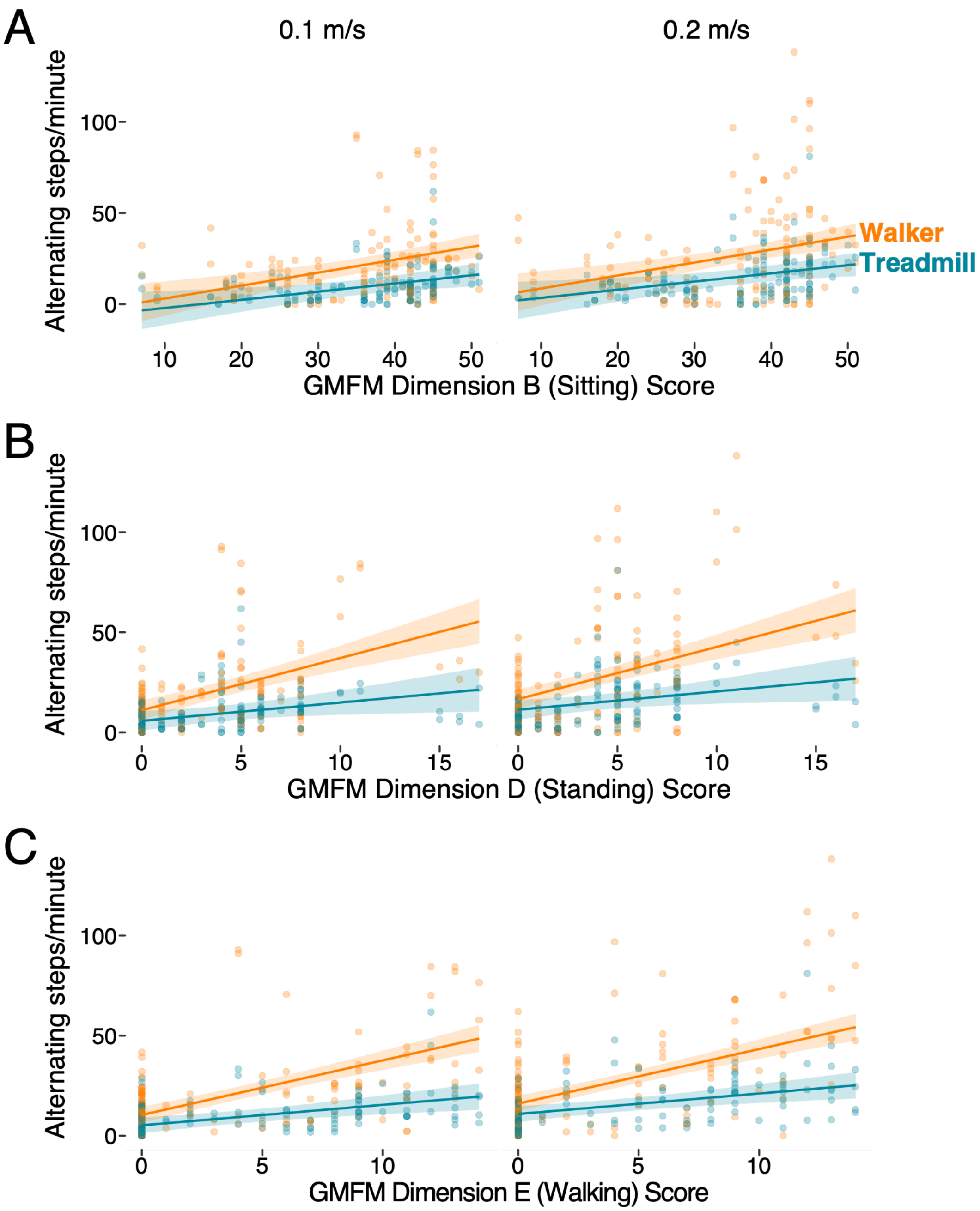


**Supplemental Figure 3**. Developmental trajectory of alternating step rate as a function of gross motor skill (x-axis) and task (colors; blue = treadmill, orange = walker): (A) Sitting skill (Dimension B of the GMFM); (B) standing skill (Dimension D of the GMFM); (C) Walking skill (Dimension E of the GMFM). Banded lines represent the model estimation with 95% confidence interval. Circular points represent trial-level individual data from each infant. Across (A), (B), and (C), the left panel includes data from the slower trials (0.1 m/s) and the right panel includes data from the faster trials (0.2 m/s). Models follow the same overall pattern as the age model reported in paper (Figure 2B and Table 2).
